# Rapid characterization of the propagation of COVID-19 in different countries

**DOI:** 10.1101/2020.06.09.20126631

**Authors:** Patricio Vargas, Sebastián Allende, Eugenio E. Vogel, Sigismund Kobe

**Author notes:** Corresponding author: Prof. Dr. Eugenio E. Vogel.

## Abstract

**BACKGROUND:** COVID-19 has spread rapidly, and there are still many characteristics of this new disease to be unveiled. We propose a simple method to calculate a “figure of merit” *F*_*C*_ to provide an early characterization of the disease status in country *C*.

**METHODS:** We use mathematical tools to adjust a Gaussian function to the daily increase of infected patients. Maximum value and full width half maximum of the Gaussian are characteristics of the development of the development of the pandemic in each country. These parameters are supplemented by the testing volume and the mortality rate to produce just one characterizing parameter: *F*_*C*_. In addition, the stability of the Gaussian fits was calculated within an entire week towards the end of the study period. Seventeen different countries were fully considered, while others are considered when discussing particular properties. Data employed is publically available.

**FINDINGS:** Fitted Gaussian functions render effective information about the development of COVID-19. The number of critical days vary between 11 (South Korea) and 52 (Mexico). *F*_*C*_ varies between 1 (Australia) and 899 (Mexico). The epidemic appears stabilized in some countries and unstable in others. Some large countries are experiencing fast development of the propagation of the disease with high *F*_*C*_. A correlation between low (high) values of the mortality rate (and to some extent *F*_*C*_) and the presence (absence) of BCG vaccination is exposed.

**INTERPRETATION:** The adjustment of a Gaussian to daily data of COVID-19 in each country reveals the different propagation dynamics, properly characterized by the parameters proposed here. Testing plays a clear role to control the spread of the disease. Mortality rate spans more than one order of magnitude and is somewhat related to permanent massive BCG vaccination. The figure of merit, *F*_*C*_, introduced here spans more than 2 orders of magnitude which makes it a useful indicator to quickly find out the status of the pandemics in each territory. Geography plays a role: low population density and isolated countries can be efficient in controlling the spread of the disease.

**HIGHLIGHTS:**

- An easy-to-evaluate parameter is defined to rapidly establish the status of the evolution of COVID-19 in any given territory. 17 countries in 4 continents are chosen to apply it and compare different evolutions.
- The parameter or figure of merit (F_C_) is dynamic: it combines testing, mortality rate, and characteristics of the Gaussian function that fits the new daily contagions.
- *F_C_* spans more than two orders of magnitude which makes it a very sensitive indicator to promptly detect second waves or local outbreaks.
- It is found that countries with massive BCG vaccination present low COVID-19 mortality rates and low values for *F_C_*.

## INTRODUCTION

There are several models that explain the evolution of a pandemic, generally based on the initial proposal of Kermack and McKendrick (Kermack and McKendrick, 1927), with additional improvements and variations through the years. Such analyses are more suitable for a posteriori discussions to prevent future outbreaks of the same or similar diseases. More recent approaches have considered the effect of quarantine, social distancing and even local heuristic behaviors to help in the control of the more recent 2003 SARS outbreak (Glasser et al., 2011).

With the current rapid spread of the new coronavirus COVID-19, indicators are needed that can promptly guide to better decision-making. At the moment we write this article there is no vaccine against this virus and is out of control in several places in the world. Moreover, new outbreaks or “a second wave” or even virus mutations are still possible so authorities need easy information to help in rapid decisions as to decide upon the best strategy for the territory, population density, habits, season of the year among several considerations. It is clear that there is no universal way of controlling the disease but learning from countries that have been successful in stopping the propagation of the disease can help to adapt local measures that are appropriate to the local population. This is the spirit of this paper: we propose a way to produce indicators or parameters that might help in this direction. Instead of providing an exhaustive mathematical analysis of the functions involved, we propose a direct and easy-to-use empirical approach to the evolution of pandemics in order to report on the characteristics of the disease in each territory. Over time, this can guide authorities in seeking timely corrections to available measures and over borders collaborations.

We are aware of the cascade of material presently published on COVID-19. Our search did not yield any results from documents published along the line we propose here.

## METHODS

### Data

There are several public databases handling live statistics for COVID-19. The Johns Hopkins University (JHU, 2020) has provided a very helpful one, with interactive maps. Another source with more emphasis on graphical analysis is provided by Worldometers (Worldometers, 2020). We will use the data continuously updated by the latter because of its tabulation and functional analysis, which is consistent with the idea of the present study. The population of each country will also be obtained from there and is given in the second column of Table I, which will summarize our main results.

**Table I.** Summary of data used in this paper and parameters reported here. Column 1: country; Column 2: population in thousand inhabitants; Column 3: Number of applied COVID-19 tests; Column 4: Test density (ratio between the last two columns); Column 5: accumulated number of positive tests; Column 6: accumulated number of deaths; Column 7: percentage of mortality (based on the ration of the two previous columns); Column 8: BCG vaccination status; Column 9: Maximum of fitting Gaussian; Column 10: FWHM of the fitting Gaussian; Column 11: Figure of merit proposed in this paper. Data from https://www.worldometers.info/: <u>columns 2, 3, 5, and 6 extracted on May 5, 2020</u>; Gaussian fits with data up to April 29, 2020, except for Brazil fitted up to April 24, 2020 due to instabilities of later data.

| Country | $P_C^*$ | $U_C$ | $T_C$ | $n_C$ | $d_C$ | $M_C$ | BCG | $A_C$ | $\Gamma_C$ | $F_C$ |
| --- | --- | --- | --- | --- | --- | --- | --- | --- | --- | --- |
| Australia | 25440 | 664756 | 26.1 | 6849 | 96 | 1.4 | 1986 | 383 | 15 | 1.2 |
| South Korea | 51269 | 640237 | 12.5 | 10804 | 254 | 2.4 | Yes | 678 | 11 | 2.7 |
| Israel | 8627 | 417100 | 48.4 | 16289 | 238 | 1.5 | 1982 | 539 | 29 | 5.5 |
| Norway | 5412 | 189657 | 35.0 | 7928 | 215 | 2.7 | Yes | 240 | 30 | 10.3 |
| Chile | 19116 | 222095 | 11.6 | 22016 | 275 | 1.3 | Yes | 482 | 47 | 12.7 |
| Austria | 9006 | 69718 | 31.7 | 15650 | 606 | 3.9 | 1990 | 718 | 19 | 18.5 |
| Germany | 83784 | 285883 | 30.4 | 166745 | 6993 | 4.2 | 1998 | 5531 | 27 | 24.6 |
| Switzerland | 8654 | 2547052 | 33.1 | 30009 | 1795 | 6.0 | 1987 | 1063 | 26 | 57.7 |
| USA | 330590 | 7681934 | 23.2 | 1235456 | 72149 | 5.8 | No | 33896 | 37 | 95.3 |
| Iran | 83993 | 519543 | 6.2 | 99970 | 6340 | 6.3 | Yes | 2449 | 37 | 110.6 |
| Spain | 46755 | 1932455 | 41.3 | 250561 | 25613 | 10.2 | 1981 | 6904 | 34 | 124.1 |
| Italy | 60462 | 2246666 | 37.2 | 213013 | 29315 | 13.8 | No | 5206 | 39 | 124.4 |
| Belgium | 11590 | 493325 | 42.3 | 50509 | 8016 | 15.9 | No | 1508 | 33 | 161.2 |
| United Kingdom | 67820 | 1383842 | 20.4 | 194990 | 29427 | 15.1 | 2005 | 5580 | 34 | 206.9 |
| Netherlands | 17135 | 235909 | 13.8 | 41087 | 5168 | 12.6 | No | 1163 | 34 | 210.8 |
| Brazil | 212258 | 339552 | 1.6 | 114715 | 7921 | 6.9 | Yes | 3854 | 48 | 376.2* |
| Mexico | 128662 | 100041 | 0.8 | 24905 | 2271 | 9.1 | Yes | 1897 | 52 | 899.1 |

From Worldometers we extract the daily number of *accumulated contagions*. This function will be referred to as *n*_*C*_*(t)* for country *C*, where *t* is the time measured in days; the latest *n*_*C*_ values are listed in the fifth column of Table I. Thus the *number of daily cases for any day t’ in the sequence* is directly given by *D*_*C*_*(t’)=n*_*C*_*(t’)−n*_*C*_*(t’−1)*.

It has been argued that the available data are not homogeneous because countries have their own testing and diagnostic systems, leaving a different number of unaccounted infected patients. We will correct this fact by taking into consideration the testing rate for each country (*T*_*C*_, fourth column in Table I): number of accumulated tests for country *C* (*U*_*C*_, third column in Table I) over the population. Consideration will be also paid to the mortality rate *M*_*C*_ (expressed as a percentage, seventh column in Table I) relating the total number of deaths (*d*_*C*_, sixth column in Table I) to the total number of cases *n*_*C*_ (fifth column).

We will extract data from Worldometers for different countries. For homogeneity all these data were those available on April 29, 2020. Seventeen countries reflecting different continents, sizes, and reactions to COVID-19 will be included in the study. For reasons of space and time, we have left out several important regions and the entire continent of Africa. However, the simple methodology will allow readers to make their own calculations for other countries. The idea is precisely to show that the approach we propose below is capable of distinguishing different realities at the intermediate stages. These countries are listed in the first column of Table I.

### Mathematical tools

The *n(t)* function (accumulated number of cases for any country) grows slowly at first but soon explodes into an exponential-like behavior (convex curve). After a few days, the reaction of society imposes limitations to the spread of the virus, and growth moderates to a point where *n(t)* becomes approximately linear: it has reached the turning or inflection point. If the measures taken by the authority are effective, this linear period ends soon, so that the growth rate of *n(t)* decreases even more and *n(t)* tends to flatten out (concave curve), asymptotically approaching its saturation value *n(t=∞)*, denoted by *Nc* for country *C*. One example can be observed in the lower panel of Fig. 1. The shape of this curve can be described mathematically by a sigmoid *Sc(t)*, given by:

**Fig. 1.**
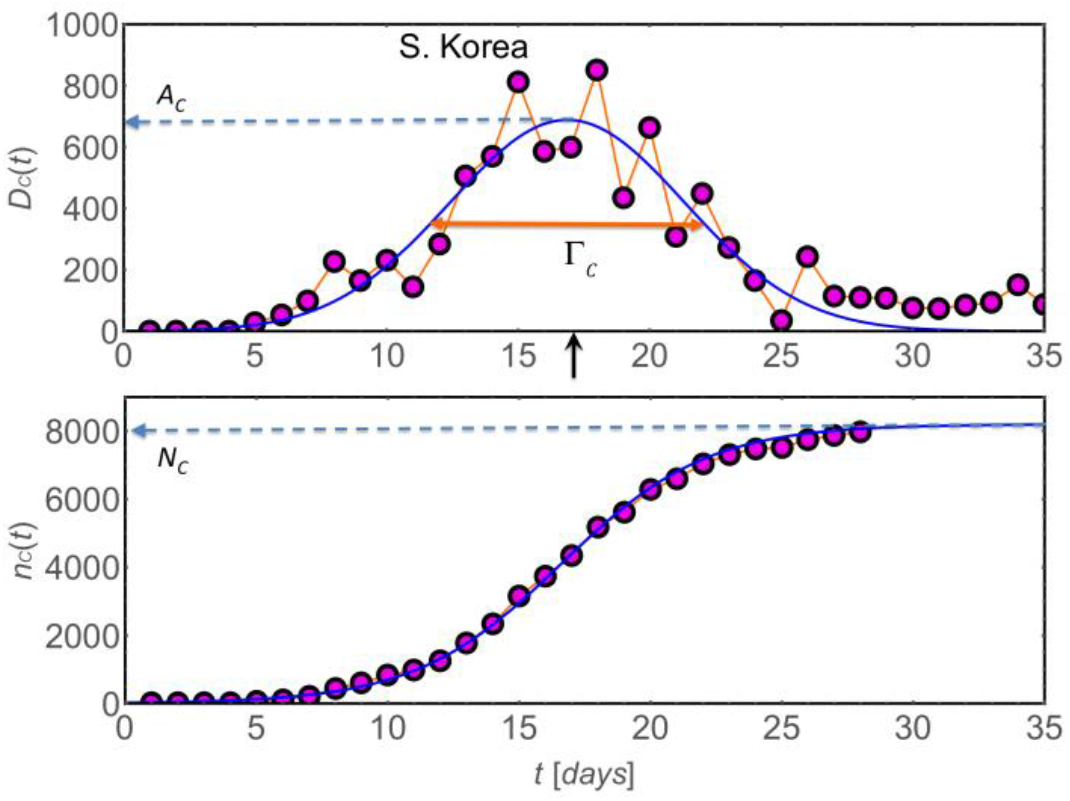
The upper panel presents the daily cases for South Korea (circles) and the adjusting Gaussian. The single dashed arrow identifies *AC*; the double arrow identifies Γ*C*, and the single vertical arrow marks *tC*. The lower panel plots *nC(t)*, the accumulated number of infected patients (circles), that smoothly lay on the curve which is the integral of the fitting Gaussian function on top *GC(t)*, without further adjustments. A possible value for *nC(∞)*, namely *NC*, is identified by a dashed arrow to the left.

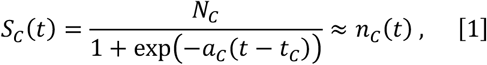

where *t*_*C*_ marks the turning point for country C, and coefficient *a*_*C*_ is related to the slope of the sigmoid of country *C* at the turning point.

The derivative of this function, *d(S*_*C*_*(t))/dt*, represents the daily cases obtained from de sigmoid whose mathematical expression can be approximately described by a Gaussian function *G*_*C*_*(t)* which can be adjusted by a least square fit to the actual daily cases *D*_*C*_*(t)* for country *C*:

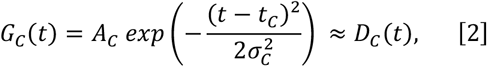

where *A*_*C*_*=a*_*C*_*N*_*C*_*/4* and *σ*_*C*_ corresponds to the standard deviation, which is related to *a*_*C*_ by the condition:

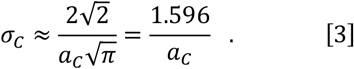

An example of this curve can be found in the upper panel of Fig. 1. The amplitude *A*_*C*_ of the Gaussian is important since it tells what is the expected number of infected people in the days of maximum crisis, a fraction of which will be demanding specialized hospital treatment. These values are given in column 9 of Table I.

Another parameter characterizing the Gaussian function is the full width half maximum (FWHM) designated by Γ_*C*_ linked to *σ*_*C*_ by the simple relationship:

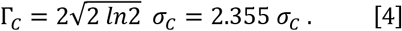

We shall use this parameter (measured in days) to characterize the duration of the critical period of the disease, which has implications on the economy of country *C*. Values of Γ_*C*_ are listed in the tenth column of Table I.

We can define a *figure of merit F*_*C*_ for any country C by combining previously defined *A*_*C*_ and *Γ*_*C*_ with other relevant parameters. The former will be normalized with respect to the population of the country *P*_*C*_.

One additional parameter is the normalized COVID-19 tests *T*_*C*_*(t)* which can be defined by the ratio of the total number of tests *U*_*C*_*(t)* performed in country C up to time *t*, over its population *P*_*C*_:

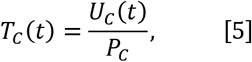

higher *T*_*C*_*(t)* values ensure better response to control de spread of the disease. Another important indicator is the mortality rate *M*_*C*_*(t)* of country *C*, defined as the percentage of deaths *d*_*C*_*(t)* with respect to the total number of positively diagnosed cases *n*_*C*_*(t)*, namely:

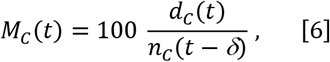

where *δ* is the average delay between the diagnosis and the death of a fatal patient. It is easy to find out the phase delay of the death curve to the diagnosis curve for any country, and we have found that although this varies a bit among countries it remains close to 9-12 days. It we use the latter value to evaluate the mortality of Germany we get 4.6 %; if *δ* is ignored (*δ*=0) the value is 4.2 %. For the purposes of the tabulation of this parameter, we will ignore *δ*, which will have a negligible effect when comparing among countries in later stages of the pandemic, since all countries will be affected by similar corrections. Lower *M*_*C*_*(t)* values point to better handling of the infected patients.

FC(t) or figure of merit can be defined as:

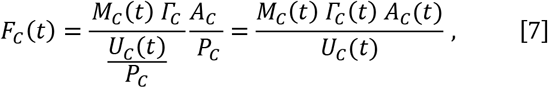

where low values of *F*_*C*_*(t)* point to better performance in handling the critical period of the disease.

This figure of merit does not mean any *a priori* judgment on the public health strategy followed by each country. It only represents the situation of a snapshot taken at a time *t* which can evolve in different ways from there on. However, A low *F*_*C*_*(t)* value can indicate that hospital facilities are expected to be enough, and/or tests are available in proportion to the population, and/or the mortality rate is low, and/or the expected duration of the crisis is short. Values of *F*_*C*_*(t)* are given orderly in the last column of Table I.

To go beyond this static image, we propose to analyze a week (usual human cyclic unit) during this study for a dynamical testing of the Gaussian fit. Then we calculated the “instantaneous” FWHM values for each country *γ*_*C*_*(t*) for the 7 days ending on April 19, 2020. This will be an indication of the trend of Γ_*C*_ both in its time derivative and in its standard deviation: large values of the latter could mean unstable status in the control of the epidemic.

### BCG Data

The Bacillus Calmette-Guérin (BCG) vaccine has been inoculated for decades worldwide. However, this has not been applied evenly to different countries. It has recently been proposed that populations on which this vaccine was massively administered at early ages are better able to resist the consequences of COVD-19. Two registered protocols for clinical trials to study the effects of BCG vaccination have been proposed (NCT04327206, 2020; NCT04328441, 2020) The World Health Organization released a scientific report on April 12, 2020, clarifying that this has not been clinically established and warning against vaccination in an attempt to stop the pandemic (WHO, 2020). Nevertheless, independent research is beginning to confirm some correlation between previous BCG vaccination and COVID-19 mitigation (Shet et al., 2020).

To contribute to this discussion, parameters in Table I will be compared with the existence of vaccination for each country as indicated in column eight of this Table. Data for BCG vaccination were obtained from a World Atlas on the subject (Zwerling et al., 2011).

## RESULTS

We have chosen the case of South Korea to illustrate the method. This is presented in Fig. 1, where the graphical parameters included in Table I and in the previous algebra are presented.

This procedure was repeated for all countries in Table I (and others not fully considered here, but used as reference in the discussions). A mosaic tracing Gaussian functions for Australia, Germany, Spain, and United Kingdom (UK) is shown in Fig. 2 for different stages of the Gaussian fit: 8/8, 7/8, 6/8, and 5/8 respectively. Obviously the quality of the information extracted from this procedure relies on the stage of this process. Moreover, local outbreaks or a general new wave can modify this description, which is nothing but a snapshot taken on April 29, 2020.

**Fig. 2.**
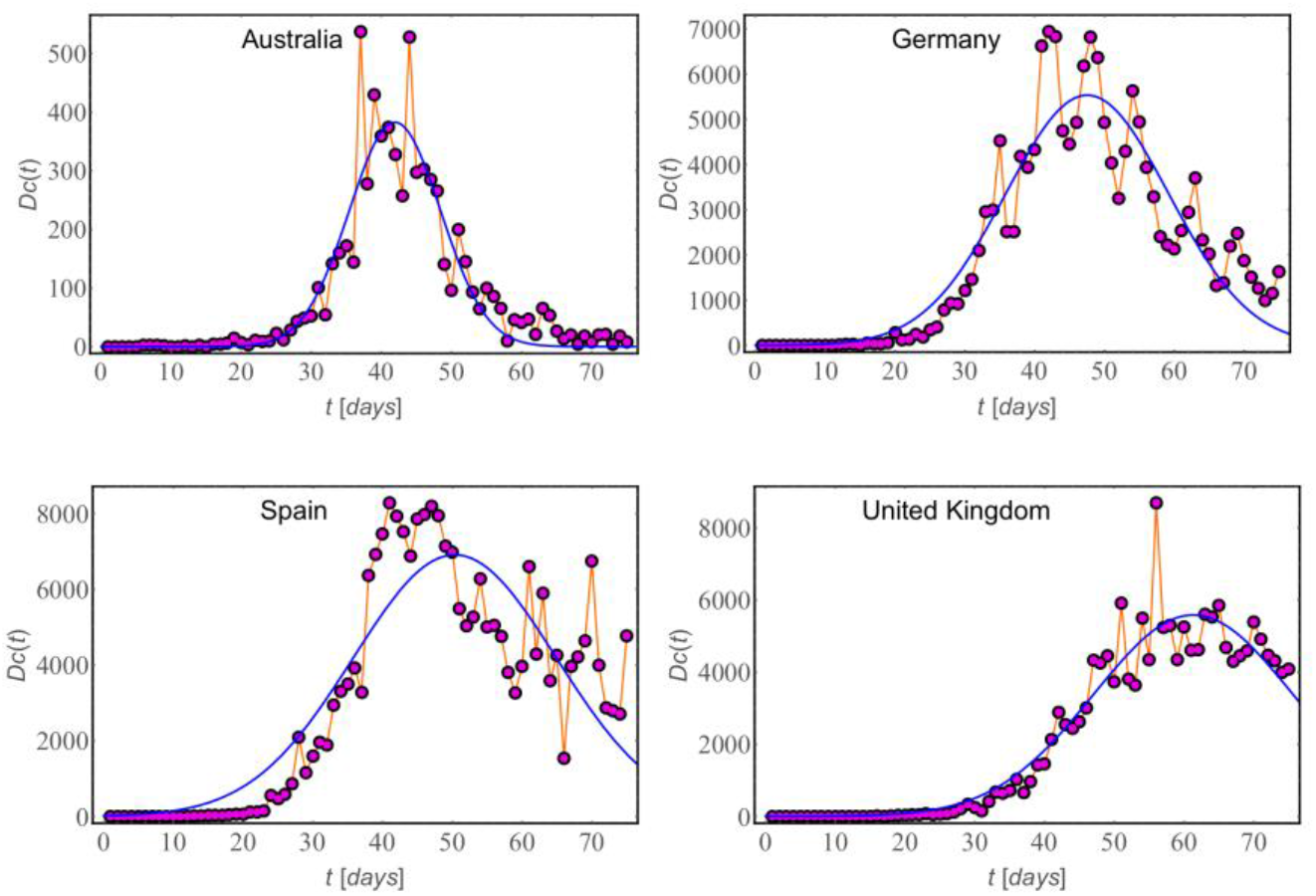
Mosaic of 4 Gaussians corresponding to 4 different status of Gaussian fit: Australia complete 8/8; Germany: almost complete 7/8; Spain: already descending 6/8, and United Kigndom going over the maximum, about 5/8.

We complemented this static analysis with dynamic calculations for the last week (7 days) ending on April 19, which is day 0, going back to April 13, which is day -6. Each of these 7 settings means a complete set of parameters. We focus here on the daily FWHM values for each country, namely *γ*_*C*_*(t)*. Fig. 3 plots the weekly average <*γ*_*C*_> for all countries in Table I, with error bars obtained from the corresponding standard deviations. The inset shows the function *γ*_*C*_*(t)* for five selected countries, with Australia already stabilized and the other tending to larger values of *γ*_*C*_*(t)*; the case of Brazil is quite different reflecting great dispersion and oscillations of the reported data.

**Fig. 3.**
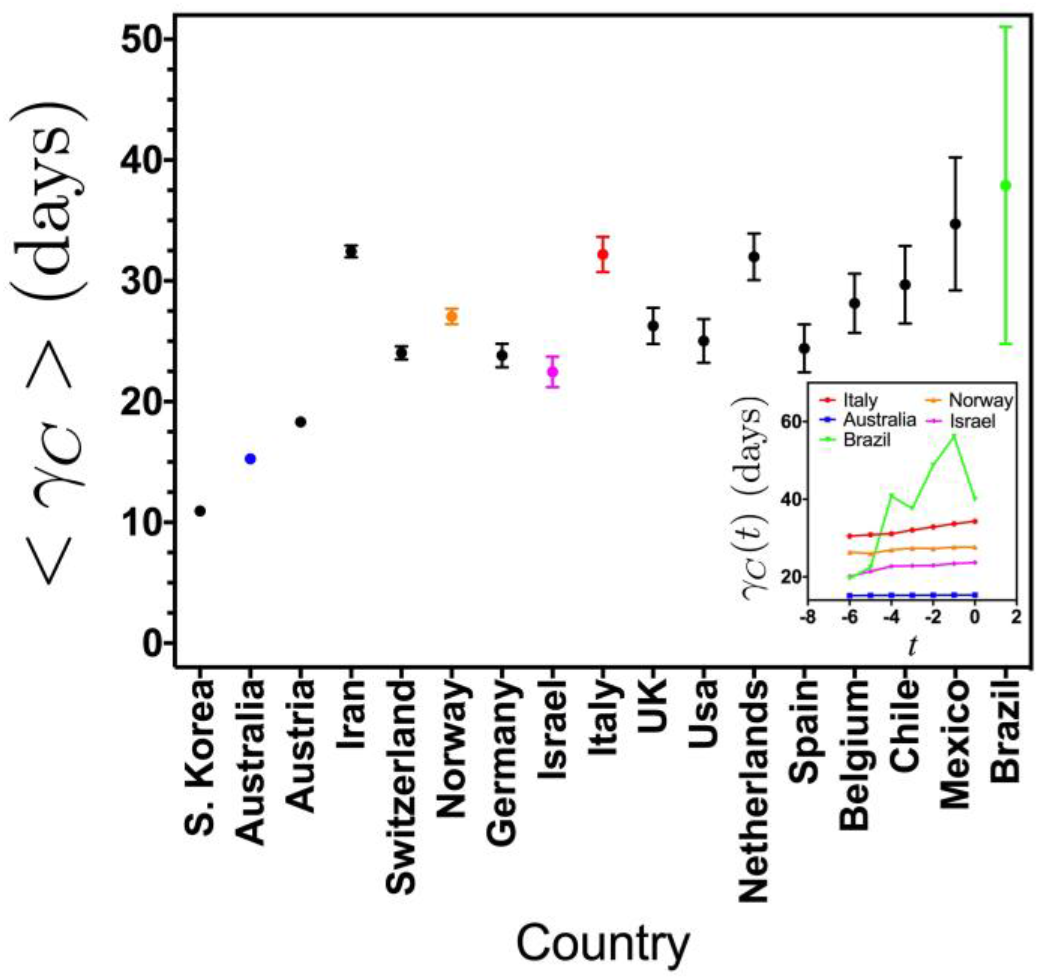
Average FWHM values with error bars for a week towards the end of this study. The insert presents functions *γC(t)* for 5 selected countries showing different behavior.

## DISCUSSION

Despite fluctuations in the daily cases of COVID-19, it is possible in general to adjust a Gaussian function to high density sets of data pointing to a possible mathematical description of the evolution of the pandemic in different countries, as shown in Figs. 1 and 2. The error bars obtained from a week in Fig. 3 indicate the stability of the data for different countries.

Table I orders the 17 countries according to increasing values of *F*_*C*_. However, this is intended just to provide a rapid indication of the status of the country at the moment this snapshot was taken (April 29, 2020). The analysis should continue to correlate this figure of merit with other relevant parameters.

It is clear that testing plays a crucial role is the prevention of any epidemic and it is also the case for COVID-19. Thus, *U*_*C*_*(t)* appears directly in the denominator of Eq. (7) and indirectly in the mortality rate *M*_*C*_*(t)*. This is the main reason to bring Brazil and Mexico to the end of the Table, but the relatively large Gaussian parameters also contribute to their high *F*_*C*_ values.

The eighth column of Table I reports on the country’s policy regarding mass vaccination for BCG (tuberculosis): “Yes” means that vaccination has been permanently imposed in this country for decades; “No” means that mass BCG inoculation was never imposed; the year indicates when mass BCG inoculation was stopped in this country. As it can be seen, countries with permanent BCG vaccination tend to have low *F*_*C*_ values, while countries where vaccination was never present or stopped a long time ago tend to have high values for the figure of merit proposed here. Norway and Switzerland have similar normalized testing *T*_*C*_ (35.0, 33.1) but a large difference in their *F*_*C*_ values: 10.3 and 57.7 respectively. It is clear that the main difference relays in the value of *A*_*C*_, namely under similar conditions these two European countries present very different contagion propagation. What makes the difference? Norway has maintained massive BCG vaccination while Switzerland stopped it in 1987. A similar comparison can be performed between Chile and The Netherlands with similar population and normalized testing, the former with massive BCG vaccination while the latter never had it. This is also evident when comparing neighboring countries with similar culture and ethnic composition: Norway (Yes for BCG vaccination) has better *F*_*C*_ than Sweden (No for BCG vaccination; not in Table I); the same comparison holds for Portugal (not in Table I) and Spain. A special situation is found for UK: vaccination was started decades after it had been in effect in continental Europe and it was also stopped after most European countries did. However, the net effect in number of years without vaccination for the presently living population is similar to other European countries.

However, the most dramatic example of a small country with no permanent BCG vaccination is Ecuador, the only one in South America unprotected from tuberculosis. Our attempts to include it in the present study failed due to the low quality of the data: *D*_*C*_*(t)* looks like a sequence of random numbers rather than the description of a phenomenon obeying natural laws. Very recently, government officials in Quito acknowledged that there was large numbers of unaccounted contagions and deaths (The Guardian, 2020).

USA has the higher *F*_*C*_ value among the countries that never had massive BCG vaccination; however, USA receives large amount of immigrants from countries where BCG vaccine is given to babies and kids, so most of the immigrants have it. The other three countries without permanent BCG vaccination occupy places at the bottom of Table I. The only country with permanent BCG vaccination found in the lower places of Table I is Iran, which has a poor testing density and high mortality rate. Although BCG inoculation does not immunize against COVID-19 it helps to control the expansion of the disease to some extent. Obviously, it is not enough: Brazil and Mexico have massive BCG vaccination, but other conditions have to be fulfilled in order to cope with the virus expansion.

Let us have a look to the upper part of the Table, with *F*_*C*_ under 30. What are the common features of these countries: Testing density *T*_*C*_ over 10.0; mortality rates *M*_*C*_ under 5.0; permanent or partial BCG vaccination; good or partial control of the daily contagions as appreciated from the Gaussian fits. At this moment there is nothing to be done with respect to BCG. However, testing can be increased, medical facilities can be increased according to the reality of each country and social restrictions can be imposed to help to control the disease. The right combination of these ingredients and other regional parameters need to be evaluated locally.

In addition, we also examined the dynamic behavior of the data over one week to find out if the Gaussian adjustments *γ*_*C*_*(t)* are already stabilized. This is shown by the error bars in Fig. 3; recent trends are seen from graphs such as those included in the inset of this figure. The abscissa carries the 17 countries in increasing order of the error bars. Although the average widths of the Gaussians are scattered around 30 days, the daily oscillations themselves can be huge. Brazil is a case on its own: the huge oscillations shown in Fig. 3 continued in the following days to the point that after April 25 was impossible to fit a Gaussian any more. That is the reason *A*_*C*_ and Γ_*C*_ for Brazil in Table I had to be calculated on April 24 to have some crude approximation for these parameters. In a sense, the huge error bar for Brazil in Fig. 3 was an announcement of the upcoming instability regime to come. Mexico and to a lower extent Chile and Belgium can fall in this oscillatory regime if no appropriate measures are taken soon.

Which is the optimal way to control COVID-19? There are two possible scenarios: A) with a vaccine against this virus, B) without such vaccine. At the moment this is a bet although there are promising news revealing progress in the development of the vaccine. Let us discuss both alternatives. A) In this case the optimal situation is a short critical period affecting the economy (small Γ_*C*_), a low value of the demand of medical facilities (small *A*_*C*_), which combine to a low value of the figure of merit *F*_*C*_. In addition, small error bars on the stabilized Gaussian functions ensure stability of this evaluation. Evidently, this requires strong measures over vast extensions of the territory to keep these indicators low all the times; the measures that lead to control the disease cannot be entirely relaxed. In the meanwhile the vaccine is produced in large quantities to inoculate all the population. Australia, South Korea, Israel, and Norway (on three different continents) have proven that this optimization is feasible. B) Without vaccine we must assume that many people will be infected sooner or later, and *N*_*C*_ will represent a substantial portion of the population. In this scenario, the only possible strategy is to try to keep *A*_*C*_ under the medical capabilities of the country upon diluting the infected cases in time meaning a large value of Γ_*C*_. From the countries listed in Table I, Spain, Italy, Belgium, United Kingdom and Netherlands present high values for the parameters considered here in coincidence with what is known as we finish writing this paper. At the moment Brazil and Mexico present unpredictable scenarios.

Countries of small population size seem to contain COVID-19 in a more efficient way. However, this is not a rule of thumb. Switzerland, The Netherlands and Belgium in spite of having populations under 18 million people, their indicators are similar to others in their neighboring countries. Thus, geography also plays a role and isolated countries like Australia and South Korea seem to have taken advantage of this fact.

Large countries without BCG massive vaccination seem to be the places where COVID-19 is harder to control. From those included in Table I we should mention USA, Spain, Italy, and UK. Of course USA stands out due to its large population (with a large portion of elderly people), diversity, and commuting networks. To get another point of view on this issue, we consider the three countries with the highest *F*_*C*_ values in Table I, looking for the number of days it took in each case to go from 1000 to 10000 contagions. After linear interpolations the results are the following: UK 11.9 days, Italy 10.5 days and Spain 12.2 days, with countries ordered by decreasing population. This evidences the fast surge that occurred in Italy: in a few days the numbers of contagions skyrocketed. In addition we can pay attention to the date the number of accumulated cases overcame 10,000 people in these countries: March 10 in Italy, March 17 in Spain, and March 26 in UK.

The propagation of COVID-19 has been from East to West and later from North to South in the Americas. Eventually, this explains the large error bars for the South American countries in Fig. 3. In this moment most of the countries in South America are on the apex of their respective fitting Gaussians, showing the strong oscillations other countries have previously experienced (See Fig. 2). We also tried to include Argentina in the present study, but the data structure is rather strange, showing days without information. Its test density is only 1.7, similar to Brazil. This is just to mention that data quality is very uneven making it difficult to analyze just one indicator so it is necessary to combine several of them.

## CONCLUSIONS

The method presented here provides a quick way to estimate the impact of COVID-19 in different countries and territories. Daily new cases can be adjusted using a Gaussian function whose FWHM (Γ_*C*_) measures the duration of the critical part of the epidemic having effects mainly on the economy. The height of the Gaussian *A*_*C*_ measures the possible response to the maximum demand of medical facilities.

Previous parameters can be combined with the testing density and the mortality rate to produce a figure of merit *F*_*C*_ with a range of values spanning over 2 orders of magnitude, providing an easy quick test for the status of the pandemic in a territory allowing to for comparisons and early detection of tendencies. An analysis based on a week of continuous data allows to find standard deviations that indicate whether the Gaussian describing each country is stabilized.

Countries with stabilized Gaussian functions and relatively low values of *F*_*C*_ show a more manageable evolution of the disease. Generally speaking, these countries have reported ongoing mass vaccination with BCG, a relationship that should be further investigated. Small isolated countries seem to be in better conditions to impose measures that effectively stop the spread of COVID-19. Controlling the spread of the pandemic up to the turning point (*t*_*C*_) seems to be possible everywhere; beyond this point progress is hard for most of the countries as revealed by the difficulty in stabilizing the Gaussian curves adjusting to the progression of the daily cases. However, some countries have succeeded to get over this curve, reporting a number of daily cases of less than 10 % the maximum of the Gaussian (*A*_*C*_), which evolves in the form of a very slow decay.

Without a vaccine against COVID-19 our only way to stop it is a global response to contain the virus, isolate it and let it die. This requires common efforts, based on rapid indicators easy to obtain at present times. Our work aims to that purpose and we believe that our parameter *F*_*C*_, eventually complemented with indicators coming from other studies, provides information to make progress in this direction. All these efforts depend on the data quality; unfortunately at present, not all the data are reliable and a huge effort should be done in defining protocols for data generation.

## Data Availability

Al data are public and properly quoted in the paper.

## Author Contributions

E. Vogel wrote the first scratch of the paper, and participated in all the subsequent discussions. P. Vargas proposed strategies for calculations and plotting curves participating in all the discussions. S. Allende got the data from the public sources and plotted curves participating in all the discussions. S. Kobe participated in the elaboration of the algebra of the present paper and participated in all discussions. All the four authors reviewed and approved the present version of the paper.

